# Human and entomological determinants of malaria transmission in the Lihir Islands of Papua New Guinea: a cross-sectional study

**DOI:** 10.1101/2024.06.12.24308686

**Authors:** Pere Millat-Martínez, Michelle Katusele, Bàrbara Baro, Bernadine Kasian, Elias Omera, Esther Jamea, Lina Lorry, Aina Casellas, Dan Ouchi, Chilaka Wali, Sylvia Raulo, Arthur Elizah, Peter Kaman, Absalom Dau, Muker Sakur, Lemen Kilepak, Siub Yabu, Nelson Koata, John Kave, Michael Toa, Christopher Urakusie, Charles Kongs, Frank Kisba, Moses Laman, Oriol Mitjà, William Pomat, Stephan Karl, Quique Bassat

## Abstract

**Background:** The Lihir Islands of Papua New Guinea, located in an area with high burden of malaria and hosting a large mining operation, offer a unique opportunity to study transmission. There, we investigated human and vector factors influencing malaria transmission.

**Methods:** In 2019, a cross-sectional study was conducted on 2,914 individuals assessing malaria prevalence through rapid diagnostic tests (RDT), microscopy, and quantitative PCR (qPCR). A logistic regression analysis identified infection-associated factors. *Anopheles* species distribution, biting behaviours, and sporozoite carriage were assessed through human landing catches and larval surveys.

**Results:** Overall malaria prevalence (any species) was 3.6% by RDT, 4.5% by microscopy, and 15.0% by qPCR. *P. vivax* accounted for 37.1% of infections, *P. falciparum* for 34.6%, *P. malariae* for 3.0%, *P. ovale* 0.2%, and mixed infections for 24.5%. Prevalence (qPCR) varied across geographic areas, from 8.5% in the mine-impacted zone (MIZ) to 27.0% in the non-MIZ. Other factors independently associated with infection risk included cohabiting with an infected individual (aOR=1.94, 95%CI: 1.56-2.42), and residing in traditional housing (aOR=1.65, 95%CI: 1.21-2.25). Children had double the infection risk compared to adults, and the use of long-lasting insecticidal-treated nets did not decrease risk of infection. *An. punctulatus* was the major vector in one of the four geographical areas; while *An. farauti* was predominant in the rest of them, both with an early biting behaviour but with different biting intensities by geographical area. Entomological inoculation rates ranged from 26.9 (95%CI: 12.3-45.2) infective bites per person-year in the MIZ to 441.3 (95%CI: 315.7-572.1) in the non-MIZ.

**Conclusions:** Malaria transmission and infection was lower in the MIZ compared to other areas, underscoring the potential of the mine company to enhance malaria control in Lihir. Measures focusing on at-risk groups, including vector-control and transmission interruption methods, could be used to reduce malaria burden, especially outside the MIZ.

**AUTHOR SUMMARY:** The Lihir Islands of Papua New Guinea host a mining operation on Aniolam, the main island, which implements specific vector control strategies in the mine-impacted zone (MIZ); whereas the remaining areas rely solely on distribution of long-lasting insecticidal-treated nets (LLIN). Our study compared transmission dynamics in the different geographic areas and demonstrated lower malaria incidence, prevalence, *Anopheles* density and biting intensities in the MIZ. Living outside the MIZ on Aniolam and on Malie Island was associated with a higher risk of infection, with transmission metrics rarely seen outside high burden countries in sub-Saharan Africa. Besides the geographic area, other risk factors for infection included younger age, cohabiting with infected individuals, and living in traditional housing; while the use of LLINs was not associated with a reduction of infection risk.

Our results highlight the need for tailored strategies based on local transmission patterns and could be beneficial for ongoing initiatives aimed at malaria control and elimination. In Lihir, there is an opportunity to invest and improve the collaboration between the mine company and public health authorities. Such a public-private partnership could enhance control efforts, including expansion of diagnostics and treatments, the vector control program and community education beyond the MIZ.

## INTRODUCTION

Progress in reducing the global malaria burden has stagnated since 2015, and the milestones proposed by the World Health Organization (WHO) have not been met (1). The WHO Western Pacific region is an example of this lack of progress, since estimates for incidence showed increases by 10% and mortality rates increases by 4% between 2015 and 2021 (2). This is largely due to a rise in the malaria burden in Papua New Guinea (PNG), as it accounts for 87% of all malaria cases and 94% of all malaria deaths in the region (3). It is estimated that 35.7% of the PNG population (approximately 3.33 million inhabitants) live in areas with high to moderate malaria risk (4). Malaria transmission in PNG exhibits geographical heterogeneity, with the northern coast and the islands regions being affected with high levels rarely found outside of Sub-Saharan Africa (5). *Plasmodium falciparum* and *P. vivax* are highly endemic in PNG, although the other two human parasite species, *P. malariae* and *P. ovale*, are also present (6). Hence, improving funding for and focusing malaria control efforts in PNG is warranted for reducing malaria transmission in the region (3).

While malaria symptomatic cases can be confirmed by microscopy or rapid diagnostic test (RDT), molecular tools such as polymerase-chain reaction (PCR) are required to confidently detect sub-clinical infections, given many infections present with sub-microscopic parasite densities (7). These sub-microscopic infections constitute a source of ongoing transmission, and are a common feature in malaria endemic areas, involving all *Plasmodium* species and individuals of all ages (8). In PNG, the 2019/2020 malaria indicator surveys showed that prevalence of microscopic parasitaemia in household surveys varied among different geographic areas, ranging from 0.1% in the highlands areas to 10.6% in the coastal areas of the country (9). When PCR is used to include sub-microscopic infections, observed prevalence of malaria parasites increases, as shown in a systematic review for *P. falciparum* infections (10). In *P. vivax* endemic settings, this difference between microscopy and PCR can be even higher. As an example, in a cross-sectional study performed in Madang Province during 2014, *P. falciparum* prevalence increased from 2.8% to 9.0% when comparing microscopy to PCR results, while *P. vivax* prevalence increased from 2.7% to 19.0% (11).

The main mosquito species transmitting malaria in PNG are members of the *Anopheles punctulatus* complex, namely *Anopheles farauti s.s*., *An. punctulatus s.s*. and *An. koliensis s.s* (12, 13), and cryptic species within this complex exist that can be hard to morphologically identify (14). Significant variation in vector abundance and infective biting rates are observed between villages, even within the same region (15). While there is no evidence of widespread pyrethroid resistance in the Anopheles populations in PNG (16), some studies suggest behavioural adaptation after the first nationwide distribution of long-lasting insecticide treated-nets (LLINs) between 2005 and 2009, resulting in a moderate shift towards an earlier peak biting in *An. punctulatus* and *An. farauti* (17). A survey conducted in 2016, confirmed that 25.5–50.8% of the vectors in the studied villages encountered human hosts in the evening, with an increased proportion of anophelines feeding outdoors (15).

The Lihir Islands, present unique characteristics to study malaria transmission and evaluate interventions. Aside of being located in one of the provinces with highest malaria transmission in PNG (>200 yearly cases per 1,000 inhabitants) (5), they host the world’s fifth largest gold mining operation on their biggest island, Aniolam. Newcrest Mining Ltd, the mine company, provides essential services to communities within the mine impacted zone (MIZ) surrounding the open-pit. In addition, a vector control program has been deployed, which includes regular larval source management in the MIZ and fogging targeting the mining camp areas. As such, since 2006, the company conducts drainage of small water bodies, and larviciding of big water bodies using *Bacillus thuringiensis israelensis* within this area. A prevalence study conducted in 2010, showed a marked reduction in malaria positive children found by microscopy in the MIZ (from 31.5% in 2006, to 5.8% in 2010) (18). In contrast, the rest of the Lihir Islands relies only on the services provided by the national malaria control program, which includes mass distribution of LLINs every 3 years to achieve universal coverage. Nevertheless, despite achieving 97%-98% coverage during the distribution campaigns, only 8.7% of households maintained the nets two years after distribution (19).

Nearly a decade later, in 2019, we conducted a study to characterize transmission, obtaining incidence data and performing prevalence and entomological surveys in the MIZ, and importantly, in the other geographic areas of the Lihir Islands. Factors associated with high malaria transmission, both in the human and the vector population, were assessed.

## METHODS

### Study setting

This study was conducted in the Lihir Islands, which are located 900 km northeast of Port Moresby and include Aniolam, Malie, Masahet and Mahur islands. A map of the Lihir Islands can be seen in S1 Fig. A population census conducted between 2018 and 2020, estimated a population of 26,528 inhabitants. The villages of the MIZ accommodate half of the Islands’ population, and there are 2,000-3,000 extra mobile workers staying at the mine housing facilities in Londolovit, in the MIZ. Most of the people staying in the MIZ are migrants from other places of PNG (57% of all inhabitants), living in permanent or makeshift houses and less in traditional houses that are more common in other areas of Lihir.

### Ethical statement and informed consent

This study was approved by the PNG Medical Research Advisory Committee (PNG-MRAC) with MRAC No.18.07. The informed consent process followed community and cultural values of PNG. Following consultation with and approval by community leaders, awareness meetings or notes (*tok saves*) were delivered in each village to explain the study. Permission for collecting malaria incidence data registered at health facilities was obtained from the health department of the local level government and from the district level government (Namatanai District, New Ireland Province, PNG).

For the cross-sectional study, individual written informed consent was obtained from all participants, or the parent or legal guardian of children below 18 years old, after explanation of the risks and potential benefits of the study. Children under the age of 18 were verbally assented. Those participants unable to read and/or write were verbally consented with a witness countersigning the consent form. For the human landing catches, individual written informed consent was obtained from all participants after explanation of the risks and potential benefits of the study, and chemoprophylaxis was offered to prevent infection from exposure to infectious mosquito bites.

### Collection of health system data

The health system in Lihir is highly fragmented. The MIZ contains the Lihir Medical Centre, run by an external health service provider contracted by the mine company. In contrast, in the non-MIZ, there are public health facilities, including a health centre, a health sub-centre and eight aid posts (S1 Fig), which are under-staffed with community health workers and some nursing officers. Passive case detection data registered between January and December of 2019 in all the health facilities (public and private) were extracted and digitalized for analysis of malaria incidence.

### Study population, data and sample collection

We conducted a cross-sectional study during October and November of 2019. We used a stratified random sampling strategy at village level. The sampling sizes of each stratum were calculated considering the data from the population census. Inside each stratum, all households were enumerated, geo-positioned and randomly selected. All individuals living in the selected households, defined as non-visitor individuals staying in that household for at least the preceding 2 weeks, and present at the moment of the survey, were eligible for recruitment to the study until the achievement of the sample size. A total of 2,914 individuals of all ages above 6 months and residing in 696 households across the 43 villages of Lihir Islands were included.

Following informed consent, demographic and clinical data, relation with head of the household, LLIN usage and mobility information were collected from all participants using Open Data Kit. Pregnancy was assessed through asking the pregnancy status to all female participants aged 16 years or older; no confirmatory test was conducted. A finger prick was performed by a health practitioner, and blood drops were collected for a malaria RDT (Malaria Pf/PAN Ag Combo RDT, Carestart™, USA), a blood slide with thin and thick smears for microscopy examination, 2 dry blood spots in filter paper, and a haemoglobin analysis with Hemocue® HB 301 analyser. A short clinical assessment, including axillary temperature measurement, spleen size assessment and history of last malaria episode, was conducted by a clinician, who also interpreted the result of the RDT. In case of a positive RDT result, antimalarial treatment with artemether/lumefantrine (plus full unsupervised course of primaquine if *P. vivax* was detected) was delivered following doses recommended by PNG guidelines (20). In case of anaemia, the participants were referred to the nearest health facility for assessment.

The laboratory procedures for the analysis of samples using light microscopy and quantitative PCR (qPCR) were standard and are described in the Supplementary methods in Supporting File S1.

### Mosquito and larval sampling

In parallel to the cross-sectional study, we conducted an entomological survey including human landing catches (HLC) and larval collection at eight sentinel sites distributed throughout Lihir: 3 villages in Aniolam MIZ, 3 villages in Aniolam non-MIZ, 1 village in Malie and 1 village in Masahet. These sites were selected after studying the environmental characteristics of Lihir Islands to represent the ecological diversity found within Aniolam, as well as in the outer Islands, and to evenly cover locations within and outside of the MIZ. For the HLC, 20-25 healthy consented volunteers were selected in each site. The inclusion criteria were those individuals, male or female, from 18 to 75 years old living in the selected site, and expressing a willingness to participate in HLC. Exclusion criteria were those participants who were not willing to give informed consent, and individuals with any apparent acute or chronic illness. The procedures for the HLC and the analyses of the mosquito biting frequency, biting time, indoor vs. outdoor biting, and the entomological inoculation rate (EIR) are described in the Supplementary methods, in Supporting File S1.

All potential larval habitats that could be found at the eight collection sites were surveyed and categorized as confirmed or potential larval habitat depending on the presence of *Anopheles spp*. larvae. Two more sites for potential larval habitats were included (one in Aniolam-MIZ and one in Aniolam non-MIZ). Density per habitat was estimated using a larval dipping method, and GPS coordinates and environmental variables were recorded. This allowed for mapping and characterizing each habitat. All larvae were reared to adults in a temporary field insectary, and were identified to species using light microscopy.

The molecular identification of Anopheles species and sporozoites from the adult mosquitoes’ samples are described in detail in the Supplementary methods, in the Supporting File S1.

### Statistical analysis

The sample size for the estimation of malaria prevalence in the general population was calculated for a precision of 1.5% and a confidence interval (CI) of 95%, with an estimated malaria prevalence of 20% based on the 2010 data (18), yielding a minimum required population to screen of 2734 participants. This calculation considered the population distribution by age and village based on a previous population census.

Data were described as absolute and relative frequencies for categorical variables. For continuous variables we described mean and standard deviation (SD) for normally distributed data and median and interquartile range (IQR) for skewed data. Chi-squared tests (or Fisher’s exact tests) and t-tests were performed to assess differences between groups for categorical and quantitative variables, respectively. Spearman’s Rank correlations were calculated to estimate the relationship between quantitative variables. Univariable and multivariable logistic regression models were used to determine the factors that were associated with PCR positivity. For the incidence, a binomial regression model was fitted to estimate the incidence risk ratio (IRR) and the Wald 95% CI; and the Mann-Kendall statistical test was employed for trend analysis to consider for seasonal patterns present in monthly incidence time series.

Mosquito collections conducted over the course of one night were considered to be equivalent to one person-night. It was assumed that the number of anopheline mosquitoes landing on a collector was equivalent to the number of mosquito bites. Biting frequency distributions over the course of the night were graphed as the mean frequency of mosquitoes sampled hourly from 6 pm to 6 am. Sporozoite rate was quantified as the proportion of PCR-tested mosquitoes that were positive for malaria parasites. Variation of sporozoite rates was assessed using Chi-squared test. Human biting rate (HBR) was quantified determining the total number of mosquitoes collected divided by the number of nights of collection and the number of collectors (i.e., the total exposure time). It was multiplied by 365.25 to obtain the HBR per person-year. 95% CI of each rate was calculated using the Wald method. EIR was estimated by multiplying the HBR and the sporozoite rate. Variation in vector composition and metrics among sampling location was tested using Chi-squared test.

A correlation analysis was performed taking into account the HBR and EIR, and the results of the prevalence and the incidence of the next month after mosquito collection, in each of the eight villages with mosquito collection. Spearman’s Rank was used for the calculation of these correlations, and 95% CI and p-values with were reported.

The computer packages STATA and R were used for the analysis of epidemiological and entomological data (21, 22). The significance level of all statistical tests was based on type I error rate of 0.05.

## RESULTS

### Study population

Demographic, geographic and clinical characteristics of participants included in the cross-sectional survey are summarized in S1 Table. Sex and age of the participants were representative of the Lihir Islands population, with 54.5% females and a predominance of younger individuals: <5 years, 12.4%; 5-14 years, 20.8%; 15-24 years, 21.1%; 25-34 years, 20.4%; 35-44 years, 11.4%; ≥45 years, 13.9%. Half of the participants (49.2%) were living in the MIZ of Aniolam, 33.5% in the rest of Aniolam (non-MIZ), 8.3% in Masahet Island, 4.9% in Mahur Island, and 4.0% in Malie Island. The type of house differed across Aniolam, with less traditional houses in the MIZ compared to the non-MIZ (9.3% vs 17.7%, p<0.001; Chi-squared test).

Of all participants, 649 (22.3%) had a history of fever in the preceding month, but only 23 (0.8%) had a documented axillary body temperature ≥37.8 °C, and 46 (1.6%) were taking an antimalarial at recruitment. Haemoglobin levels were below 12 g/dL in 70.5% of the participants and increased with age, with a mean of 9.4 g/dL in under 5 years-old, 10.5 g/dL in 5-14 years-old, and >11.0 g/dL in all groups older than 15 years-old (p<0.0001; Chi-squared test). Haemoglobin <8.0 g/dL was detected in 6.8% of the individuals. Pregnant women had lower haemoglobin levels than their non-pregnant child-bearing age counterparts (9.6 g/dL vs 10.6 g/dL, p=0.0005; t-test). On the other hand, 71 (4.8%) participants had splenomegaly. which differed with age, being 7.9% in participants under 5 years-old, 7.5% in 5-14 years-old, and <4.0% in all age groups older than 15 years-old (p=0.0090; Chi-squared test).

Splenomegaly was also more frequent in individuals living in the non-MIZ of Aniolam (7.3%) compared to the other geographic areas, where prevalence of splenomegaly was below 4% (p=0.0207; Chi-squared test).

Finally, 37.2% of the participants reported having slept under a LLIN the previous night. LLIN use was more common among females compared to males (41.3% vs 32.3%, p<0.0001; Chi-squared test), while sleeping outdoors was more frequent in males compared to females (6.7% vs 2.9%, p<0.0001). LLIN use also differed across age (p<0.0001), with children under 5 years-old being the ones using LLIN the most (49.3%). When asked on prevention measures to avoid acquiring malaria, less than half of the population (39.6%) responded that sleeping under LLIN prevents malaria.

### Malaria prevalence in the Lihir Islands

Overall, 105 participants (3.6%) had a positive malaria RDT, 132 (4.5%) had blood-stage parasites of any *Plasmodium* species detectable by light microscopy, and 437 (15.0%) had a *Plasmodium* positive qPCR.

RDT showed in 91 (86.7%) participants a single line for histidine-rich protein 2 (HRP2), indicating *P. falciparum* mono-infection; in 14 (13.3%) participants it showed a single line for *Plasmodium* lactate dehydrogenase (pLDH), indicating non-*P. falciparum* species infection (mainly *P. vivax* in this setting); and there were no RDTs with both lines positive (indicating *P. falciparum* or mixed infection). In contrast, microscopy showed 67 (50.8%) cases of *P. falciparum* infection, 51 (38.6%) of *P. vivax*, 7 (5.3%) of *P. malariae*, and 7 (5.3%) mixed infections (*P. falciparum* and *P. vivax)*. No *P. ovale* infection was detected by microscopy.

Gametocytes were detected in 23 (31.1%) of all positive *P. falciparum* samples, with a median (IQR) concentration of 164 (45-1,854) parasites/µL; and in 12 (20.7%) of all positive *P. vivax* samples, at 70 (28-229) parasites/µL. Gametocyte prevalence considering both species was 1.2% of the overall study population. On the other hand, median asexual parasitaemia was 442 (147-5,313) parasites/µL in any *P. falciparum* positive sample, and 164 (78-853) parasites/µL in any *P. vivax* positive sample. Aiming to compare with a previous prevalence study performed in Aniolam (18), we quantified the positive cases found by microscopy in children under 5 years-old, which were 5.0% in the MIZ of Aniolam and 19.9% in the non-MIZ (p <0.0001).

Furthermore, from the 2,908 samples with DNA material for qPCR analysis, species differentiation showed 162 (37.1%) cases of *P. vivax* mono-infection, 151 (34.6%) of *P. falciparum* mono-infection, 13 (3.0%) of *P. malariae* mono-infection, 1 (0.2 %) of *P. ovale* mono-infection, and 107 (24.5%) of mixed infection. Mixed infections included mostly the main two species, with 83 cases positive for *P. falciparum* and *P. vivax*, 15 for *P. falciparum* and *P. malariae*, 2 for *P. falciparum* and *P. ovale*, and 2 for *P. vivax* and *P. malariae*. In addition, there were 2 cases of a triple combination of species, which included *P. falciparum, P. vivax* and *P. malariae*; and 1 infection that included the four species. Positive cases for *P. falciparum*, *P. vivax* or mixed infections detected by qPCR compared with microscopy and age distribution are shown in Fig 1.

**Fig 1.**
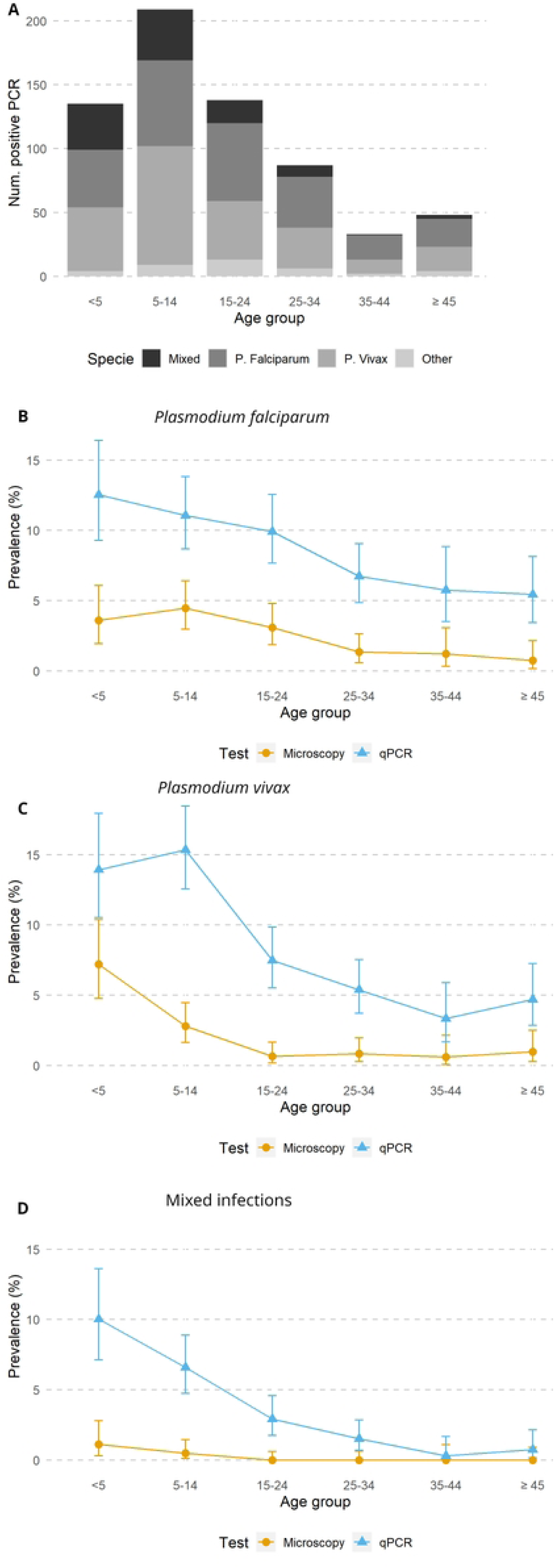
*Plasmodium* species detected in the cross-sectional survey. (A) Number of *Plasmodium* infections detected by qPCR classified by age group and species; (B) Prevalence of *P. falciparum* infections by age groups (light microscopy and qPCR); (C) Prevalence of *P. vivax* infections by age groups (light microscopy and qPCR); (D) Prevalence of mixed infections by age groups (light microscopy and qPCR). Prevalence data are shown in percentage and 95% confidence intervals.

### Factors associated with malaria infection

Associations of the population characteristics with prevalence of *Plasmodium* infection by qPCR are shown in Table 1. Prevalence was higher in males compared to females (17.6% vs 12.9%, p=0.0004; Chi-squared test), and also varied across age groups, with higher prevalence in children younger than 15 years-old (p<0.0001). On the other hand, malaria prevalence was higher in those participants with lower haemoglobin levels (p=0.0197). Malaria prevalence by qPCR was also higher in those participants with splenomegaly compared to those without (23.9% vs 13.1%, p=0.0095), and in pregnant women compared to the non-pregnant women at childbearing age (19.0% vs 10.9%, p= 0.0493). On the other hand, presenting a body temperature of ≥37.8 °C was not associated with a higher malaria prevalence (p=0.5623; Fisher exact test).

**Table 1.**
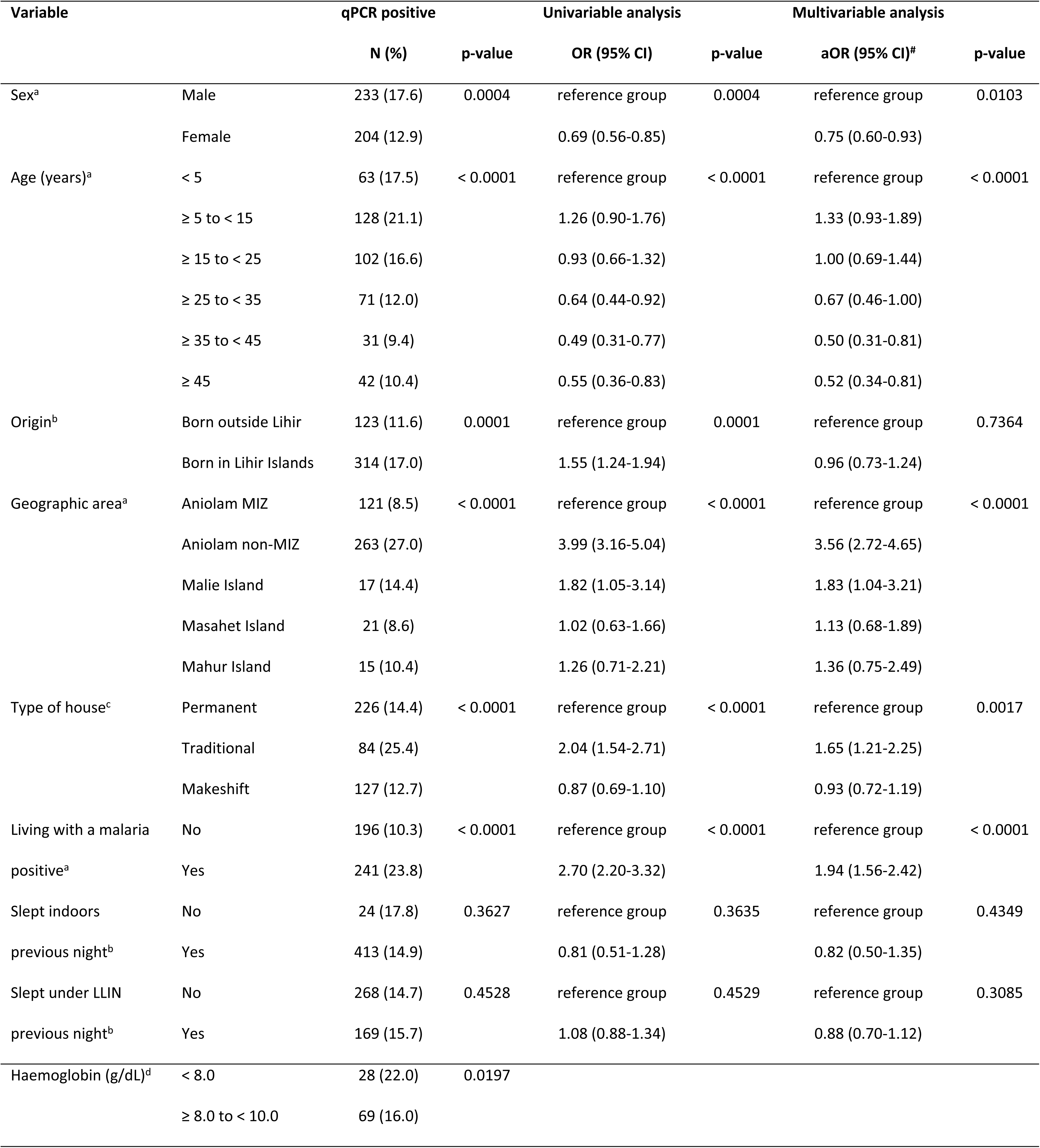
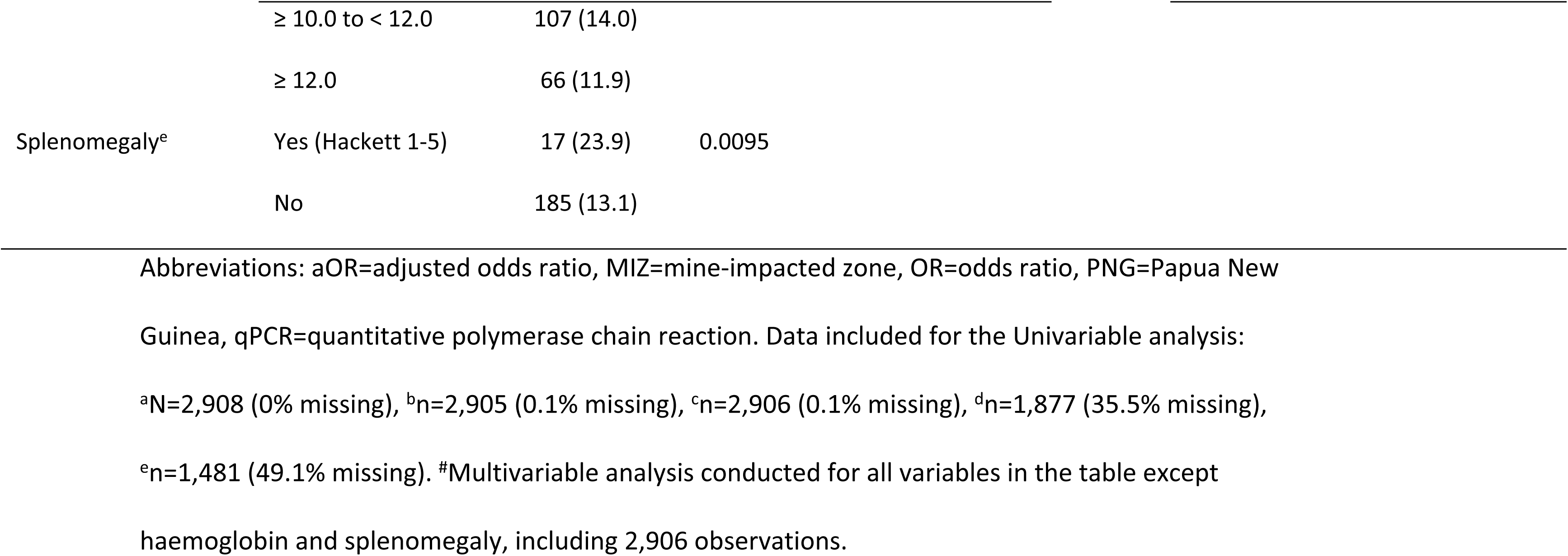
Variables associations with *Plasmodium* infection by qPCR and logistic regression models for risk factors.

| Variable |  | qPCR positive |  | Univariable analysis |  | Multivariable analysis |  |
| --- | --- | --- | --- | --- | --- | --- | --- |
|  |  | N (%) | p-value | OR (95% CI) | p-value | aOR (95% CI)<sup>#</sup> | p-value |
| Sex<sup>a</sup> | Male | 233 (17.6) | 0.0004 | reference group | 0.0004 | reference group | 0.0103 |
|  | Female | 204 (12.9) |  | 0.69 (0.56-0.85) |  | 0.75 (0.60-0.93) |  |
| Age (years)<sup>a</sup> | < 5 | 63 (17.5) | < 0.0001 | reference group | < 0.0001 | reference group | < 0.0001 |
|  | ≥ 5 to < 15 | 128 (21.1) |  | 1.26 (0.90-1.76) |  | 1.33 (0.93-1.89) |  |
|  | ≥ 15 to < 25 | 102 (16.6) |  | 0.93 (0.66-1.32) |  | 1.00 (0.69-1.44) |  |
|  | ≥ 25 to < 35 | 71 (12.0) |  | 0.64 (0.44-0.92) |  | 0.67 (0.46-1.00) |  |
|  | ≥ 35 to < 45 | 31 (9.4) |  | 0.49 (0.31-0.77) |  | 0.50 (0.31-0.81) |  |
|  | ≥ 45 | 42 (10.4) |  | 0.55 (0.36-0.83) |  | 0.52 (0.34-0.81) |  |
| Origin<sup>b</sup> | Born outside Lihir | 123 (11.6) | 0.0001 | reference group | 0.0001 | reference group | 0.7364 |
|  | Born in Lihir Islands | 314 (17.0) |  | 1.55 (1.24-1.94) |  | 0.96 (0.73-1.24) |  |
| Geographic area<sup>a</sup> | Aniolam MIZ | 121 (8.5) | < 0.0001 | reference group | < 0.0001 | reference group | < 0.0001 |
|  | Aniolam non-MIZ | 263 (27.0) |  | 3.99 (3.16-5.04) |  | 3.56 (2.72-4.65) |  |
|  | Malie Island | 17 (14.4) |  | 1.82 (1.05-3.14) |  | 1.83 (1.04-3.21) |  |
|  | Masahet Island | 21 (8.6) |  | 1.02 (0.63-1.66) |  | 1.13 (0.68-1.89) |  |
|  | Mahur Island | 15 (10.4) |  | 1.26 (0.71-2.21) |  | 1.36 (0.75-2.49) |  |
| Type of house<sup>c</sup> | Permanent | 226 (14.4) | < 0.0001 | reference group | < 0.0001 | reference group | 0.0017 |
|  | Traditional | 84 (25.4) |  | 2.04 (1.54-2.71) |  | 1.65 (1.21-2.25) |  |
|  | Makeshift | 127 (12.7) |  | 0.87 (0.69-1.10) |  | 0.93 (0.72-1.19) |  |
| Living with a malaria positive<sup>a</sup> | No | 196 (10.3) | < 0.0001 | reference group | < 0.0001 | reference group | < 0.0001 |
|  | Yes | 241 (23.8) |  | 2.70 (2.20-3.32) |  | 1.94 (1.56-2.42) |  |
| Slept indoors previous night<sup>b</sup> | No | 24 (17.8) | 0.3627 | reference group | 0.3635 | reference group | 0.4349 |
|  | Yes | 413 (14.9) |  | 0.81 (0.51-1.28) |  | 0.82 (0.50-1.35) |  |
| Slept under LLIN previous night<sup>b</sup> | No | 268 (14.7) | 0.4528 | reference group | 0.4529 | reference group | 0.3085 |
|  | Yes | 169 (15.7) |  | 1.08 (0.88-1.34) |  | 0.88 (0.70-1.12) |  |
| Haemoglobin (g/dL)<sup>d</sup> | < 8.0 | 28 (22.0) | 0.0197 |  |  |  |  |
|  | ≥ 8.0 to < 10.0 | 69 (16.0) |  |  |  |  |  |

|  |  |  |  |
| --- | --- | --- | --- |
|  | ≥ 10.0 to < 12.0 | 107 (14.0) |  |
|  | ≥ 12.0 | 66 (11.9) |  |
| Splenomegaly<sup>e</sup> | Yes (Hackett 1-5) | 17 (23.9) | 0.0095 |
|  | No | 185 (13.1) |  |

Abbreviations: aOR=adjusted odds ratio, MIZ=mine-impacted zone, OR=odds ratio, PNG=Papua New Guinea, qPCR=quantitative polymerase chain reaction. Data included for the Univariable analysis: <sup>a</sup>N=2,908 (0% missing), <sup>b</sup>n=2,905 (0.1% missing), <sup>c</sup>n=2,906 (0.1% missing), <sup>d</sup>n=1,877 (35.5% missing), <sup>e</sup>n=1,481 (49.1% missing). #Multivariable analysis conducted for all variables in the table except haemoglobin and splenomegaly, including 2,906 observations.

Regarding the geographic distribution, prevalence of both microscopic and sub-microscopic infections varied across areas (p<0.0001). The lowest prevalence (qPCR) was found in the MIZ of Aniolam (8.5%) and in Masahet Island (8.6%), followed by Mahur Island (10.4%), Malie Island (14.4%) and the non-MIZ of Aniolam (27.0%). Of note, we found highly heterogeneous malaria prevalence across villages and administrative divisions, especially in the non-MIZ, ranging from 12.4% in the south-western coast to 38.9% in the north-western coast of Aniolam. Distribution of positive cases by qPCR across all administrative divisions is shown in Fig 2A.

**Fig 2.**
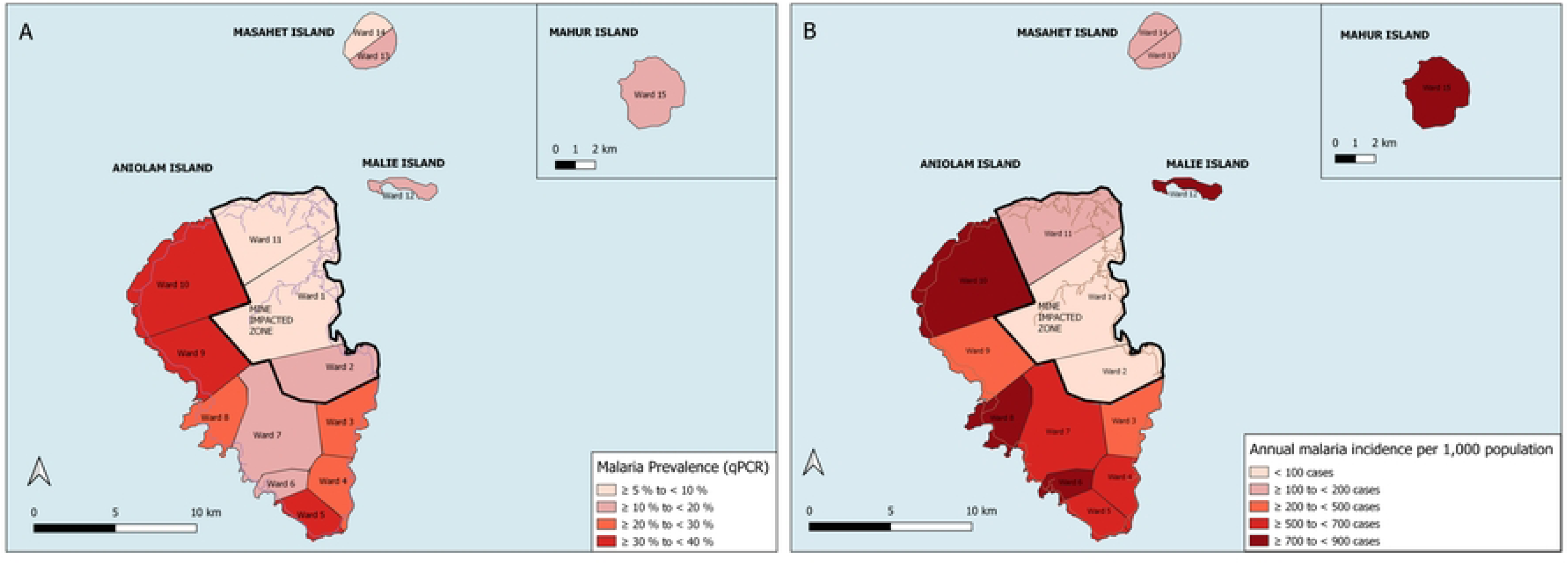
Malaria prevalence and incidence in each geographic area of the Lihir Islands. (A) Malaria prevalence by qPCR presented in percentages, with the denominator being the number of participants included in each area; (B) Incidence rates presented in annual cases per 1,000 inhabitants, with the denominator being the total population of each area (population census conducted in 2018-2020). In both maps, areas are divided by wards (administrative divisions), and the mine impacted zone comprises wards 1, 2 and 11.

We performed uni– and multivariable logistic regression analyses to identify the risk associated with being positive for *Plasmodium* parasites by qPCR (Table 1). Haemoglobin levels, splenomegaly, and presence of fever were excluded of the analysis. Females had a decreased risk of infection, with an OR of 0.69 (95% CI: 0.56-0.85). Adults above 25 years-old also had a decreased risk of infection [OR = 0.64 (95% CI: 0.44-0.92) for those between 25 and 34 years-old; OR = 0.49 (95% CI: 0.31-0.77) for those between 35 and 44 years-old; and OR = 0.55 (95% CI: 0.36-0.83) for ≥ 45 years-old individuals]. On the other hand, living with at least one other person infected by malaria increased the risk of infection, with an OR of 2.7 (95% CI: 2.20-3.32). We found an increased risk of infection for individuals living in the non-MIZ of Aniolam [OR=3.99 (95% CI: 3.16-5.04)] and for those living in Malie Island [OR=1.82 (95% CI: 1.05-3.14)]. Participants born in Lihir Islands were also more frequently infected [OR = 1.55 (95% CI: 1.24-1.94)], as well as those living in traditional houses [OR = 2.04 (95% CI: 1.54-2.71)]. Individuals that had slept indoors or had slept under LLIN the preceding night did not appear to show a decreased risk of malaria infection.

In the multivariable analysis, the strongest independent risk factor for carrying *Plasmodium* parasites was the geographic area, with the highest risk in the non-MIZ of Aniolam, presenting an adjusted OR (aOR) of 3.56 (95% CI: 2.72-4.65). Sex was also independently associated with parasite prevalence, with an aOR of 0.75 (95% CI: 0.60-0.93) in females. Risk of malaria infection also decreased with age older than 35 years-old (p<0.0001). Finally, other independent risk factors associated with parasite prevalence by qPCR were living with a malaria positive individual [aOR = 1.94 (95 % CI: 1.56-2.42; p < 0.0001] and living in a traditional type of house [aOR = 1.65 (95% CI: 1.21-2.25; p = 0.0017].

### Passive case detection and malaria incidence in the Lihir Islands

In 2019, a total of 18,419 patients were attended in the Lihirian health facilities with suspected malaria, of whom 9,207 (50%) were considered positive. Of the positive, 7,774 (84.4%) were diagnosed by RDT, 1,377 (15.0%) by microscopy, and 56 (0.6%) based solely on clinical grounds, without using diagnostic tools. Considering only the confirmed cases, malaria incidence in Lihir was 345 cases per 1,000 inhabitants, with an annual blood examination rate of 69.2%.

Incidence was similar across months (p=0.054; Mann-Kendall test), with a mean (±SD) of 767.3 (±169.7) cases per month. Similar to parasite prevalence, incidence varied substantially by geographic areas (p<0.001; Wald test, Fig 2B). In addition, incidence varied across age (p<0.001), with 53.6% of malaria cases occurring in children below 15 years-old (S2A Fig). Incidence rates and incidence risk ratios between the different geographic areas and age groups is shown in S2 Table. Incidence of each *Plasmodium spp.* varied depending on the diagnostic tool used. A comparison of diagnosed *Plasmodium* species by microscopy and RDT is shown in S2B Fig.

### Larval habitats surveillance

A total of 976 potential larval habitats were surveyed at ten entomological sites (i.e., around the same eight villages used for adult mosquito population, plus two extra villages). Overall, 92 (9.4%) of surveyed habitats were positive for *Anopheles* larvae. The distribution of the larval habitats surveyed and those positive for *Anopheles* larvae is shown in Fig 3A. The proportion of habitats positive for *Anopheles* larvae varied across geographic areas (p<0.0001; Chi-squared test), with Malie Island having the highest (29.5%), followed by the non-MIZ of Aniolam (14.2%), the MIZ (5.9%) and Masahet Island (3.9%), corresponding well with the observed abundance of adult *Anopheles* mosquitoes. The most frequent habitat for *Anopheles* larvae was the permanent groundwater (e.g., swamps), with 25% of the sampled wells positive for *Anopheles* species, followed by transient puddles (16.7%), and forest swamps (15.4%)*. Anopheles* species present in each surveyed habitat are shown in S3 Table.

**Fig 3.**
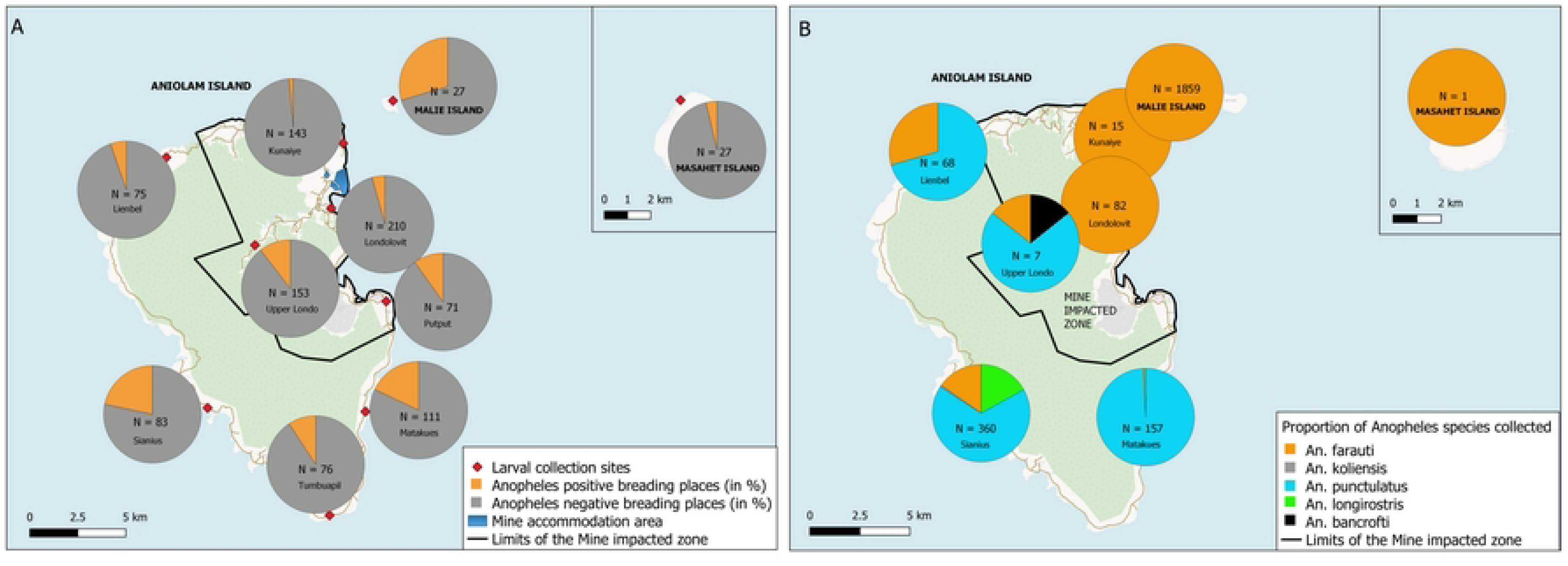
*Anopheles* species and larval habitats in the Lihir Islands. (A) proportion of *Anopheles* positive larval habitats after entomological identification of larvae grown into adults in the insectary, the number of total potential habitats surveyed and the name of each collection site is described in each chart; (B) proportions of each species of *Anopheles* collected in human landing catches measured by qPCR, the number of total *Anopheles* collected per site and the name of each collection site is described in each chart.

### Adult vector indicators

A total of 2,549 *Anopheles* mosquitoes were collected with HLCs at the eight villages and over 7,821 person-hours of collection. Of these, 2,034 (79.8%) were *An. farauti,* 448 (17.5%) were *An. punctulatus,* 61 (2.4%) were *An. longirostris*, 2 (0.08%) were *An. koliensis,* and 1 (0.04%) was *An. bancroftii*. There were 3 (0.1%) *Anopheles* mosquitoes collected for which the species could not be ascertained morphologically.

Vector species across the eight HLC collection villages are shown in Fig 3B. Only 1 anopheline specimen was collected in Masahet (*An. farauti). S*pecies distribution differed across geographic areas (p<0.0001), with *An. farauti* being the predominant vector in the MIZ of Aniolam (94.2%, 95% CI: 88.0-97.3), in Malie Island (100%, 95% CI: 89.9-100.0), and in Masahet Island (n=1). In contrast, *An. punctulatus* was the predominant species in the non-MIZ of Aniolam (75.7%, 95% CI: 72.1-79.0), followed by *An. farauti* (13.0%, 95% CI: 10.5-16.0) and *An. longirostris* (10.4%, 95% CI: 8.2-13.2).

Human biting rates, sporozoite rates, and the resulting entomological inoculation rates are shown in detail in Table 2. The HBR amongst all *Anopheles* collected (any species) varied significantly across areas (p<0.0001; Chi-squared test), with a mean of 8,487.5 bites per person-year in Malie Island, 896.4 bites per person-year in the non-MIZ, and 175.9 bites per person-year in the MIZ of Aniolam.

**Table 2.**
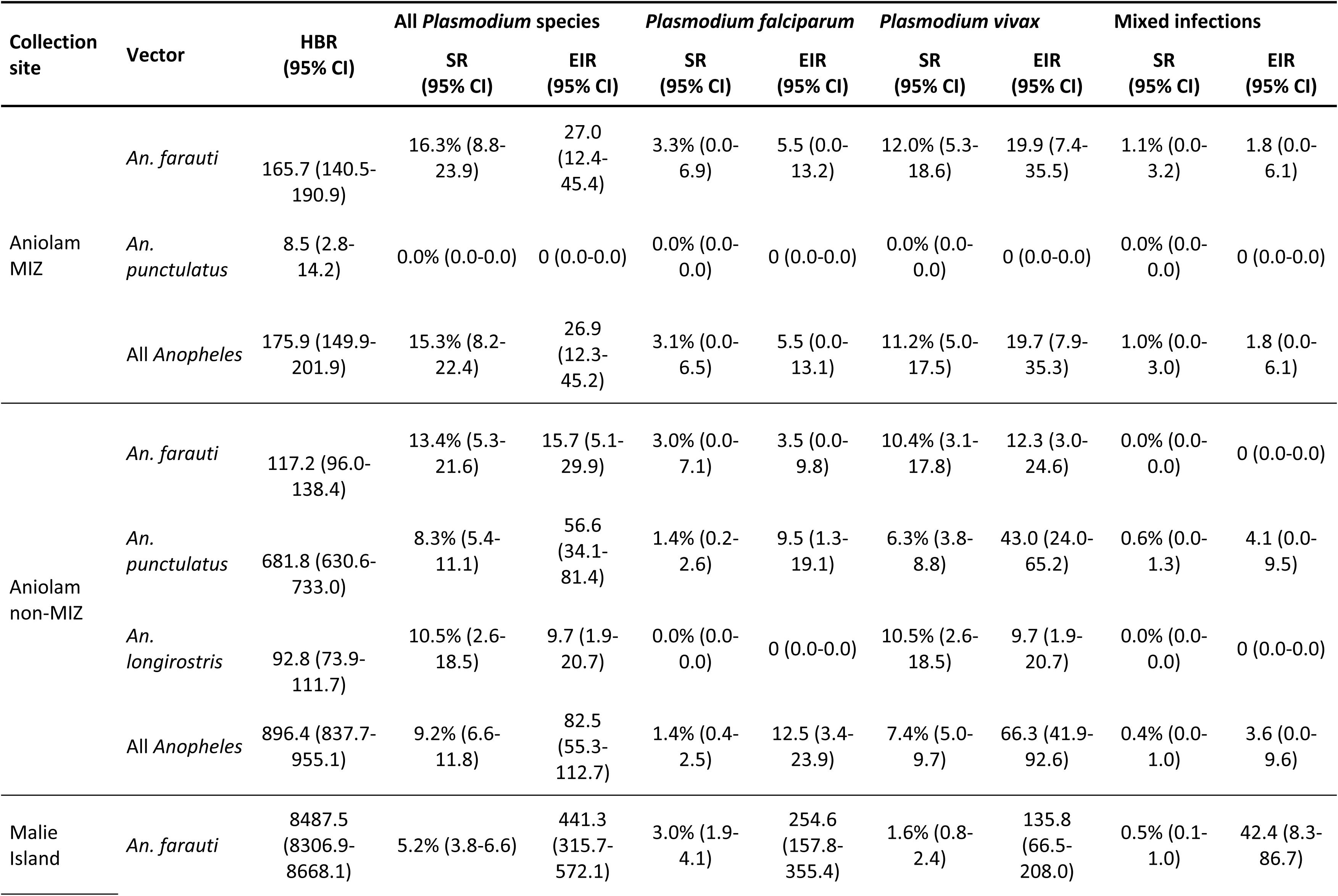
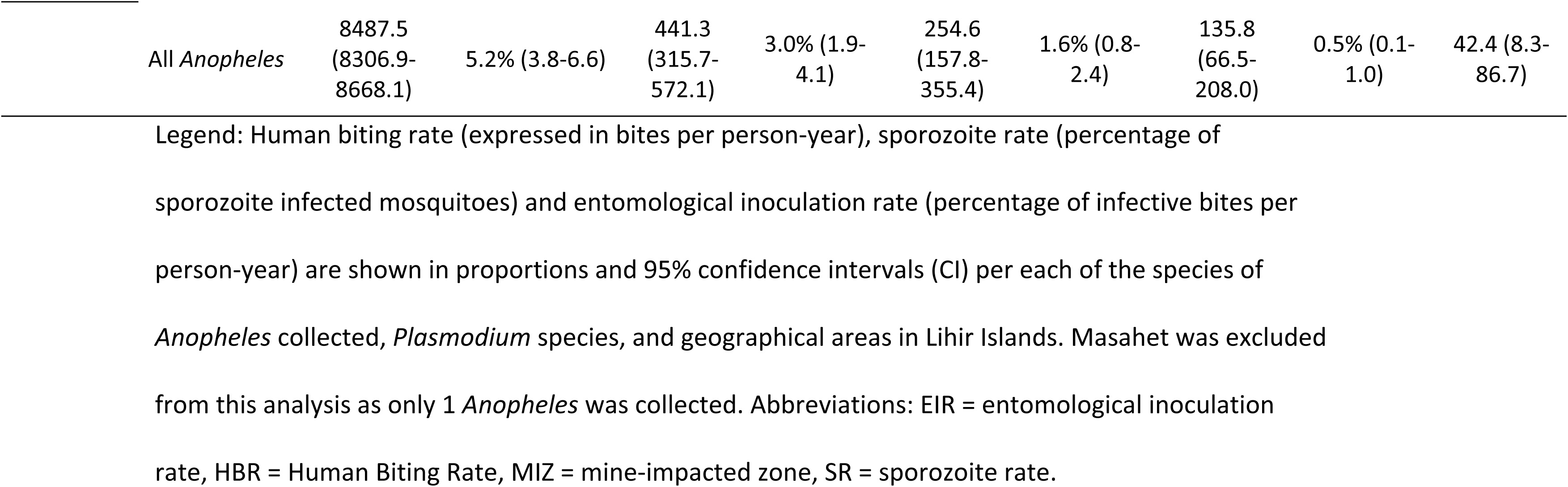
Summary of adult vector indicators in each geographic area.

A total of 1,511 *Anopheles* mosquitoes were tested for *Plasmodium* species by qPCR, which included 1,083 *An. farauti* (53% of all *An. farauti* collected), 368 *An. punctulatus* (82% of all *An. punctulatus* collected), 57 *An. longirostris* (93% of all *An. longirostris* collected), and 2 *An. koliensis* (100% of all *An. koliensis* collected). Of these, a total of 108 (7.1%) mosquitoes were positive for *Plasmodium spp* sporozoites. Overall, the sporozoite rate was 4.1% (95% CI: 3.1-5.1) for *P. vivax*, 2.5% (95% CI: 1.7-3.3) for *P. falciparum,* and 0.5% (95% CI: 0.0-0.9) for mixed infections containing both *Plasmodium* species. The vastly different biting rates within the three geographical regions surveyed resulted in very different EIR estimates, with Malie Island exhibited the highest EIR estimates, followed by the non MIZ of Aniolam, and the MIZ having the lowest EIR estimates (Table 2).

Only 26.3% of mosquitoes were captured indoors despite balanced collection effort indoors and outdoors, with *An. farauti* (26.5%), *An. punctulatus* s.s. (25%), and *An. longirostris* (26.2%) all exhibiting similar preference for outdoor biting. There was no apparent difference in the preference for outdoor biting in the different geographical zones.

Biting behaviour over the course of the night was only analysed for the two most abundant *Anopheles* species, *An. farauti and An. punctulatus* s.s, as shown in Fig 4. *An. farauti* on Malie Island exhibited an early biting behaviour with a peak at 6-7pm. *An. punctulatus*, most abundant in the non-MIZ of Aniolam, exhibited the typical late-biting profile characteristic for this species. The very low abundance of either *Anophele*s species in the other zones precluded an accurate determination of their biting profiles in these zones.

**Fig 4.**
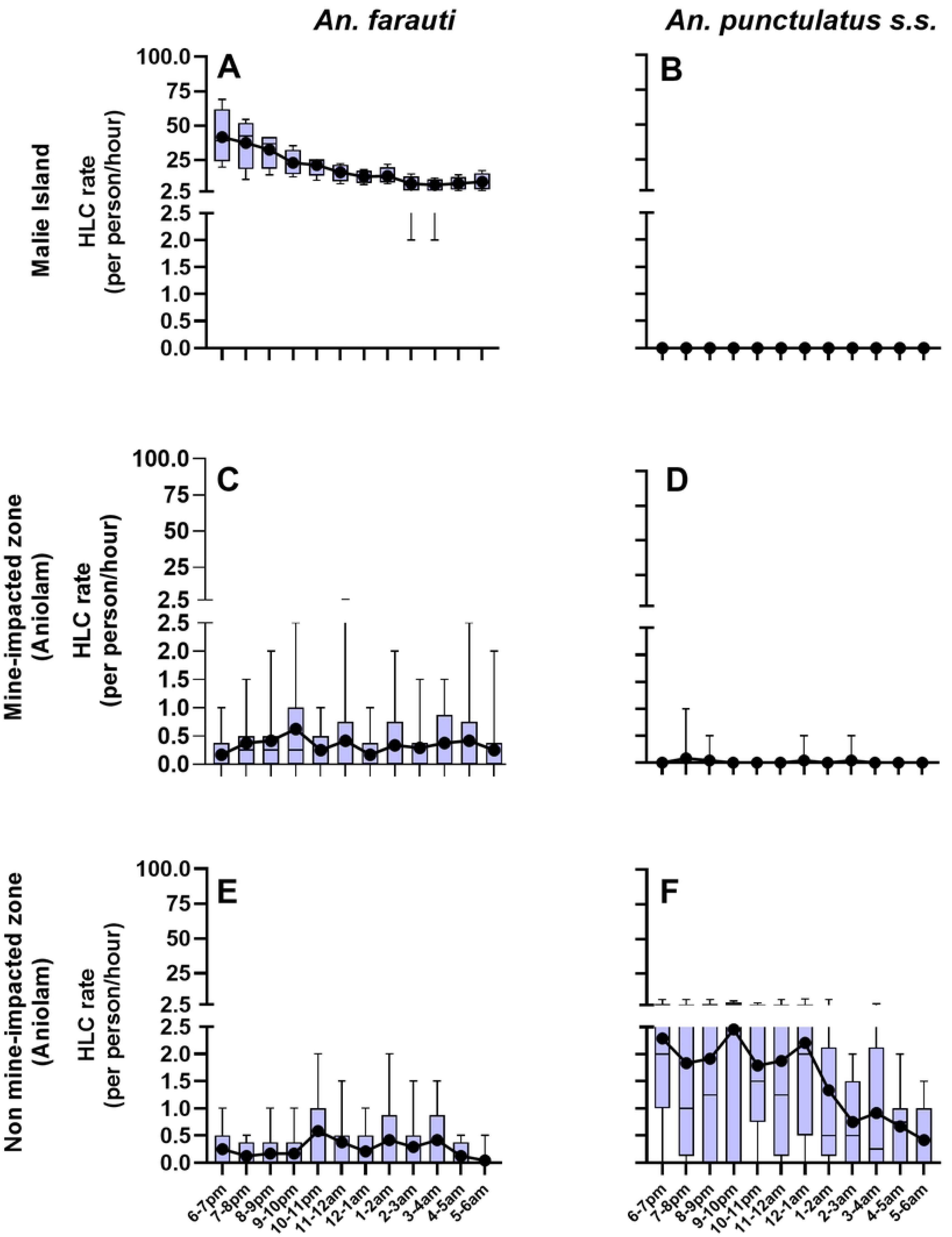
Human landing catch rates for the major vectors found in the Lihir Islands. The graphs use combined indoor and outdoor HLC rates for each night of collection per zone (Malie Island: 4 nights; MIZ and non-MIZ of Aniolam: 12 nights). (A) and (B), Malie Island; (C) and (D), the Mine impacted zone (MIZ) of Aniolam; (E) and (F), Aniolam out of the Mine impacted zone (Aniolam non-MIZ). The box and whisker plots present the median, interquartile ranges and ranges of the catch rates. The black dots represent the mean of the catch rates. Both statistics are presented to account for the highly skewed nature of the data.

### Correlation between vector and human indicators

We assessed the relationship between HBR and EIR with malaria prevalence in each of the villages where both entomological and human indicators were collected; as well as the relationship between EIR and incidence data from the month after the collection. Data on human and vector indicators at village level are shown in Table 3. Although not statistically significant, there was a clear trend towards a positive correlation between the HBR and the incidence (rho = 0.63, 95% CI: –0.15,0.93; Spearman test), as well as between the EIR and the incidence (rho = 0.52, 95% CI: –0.32,0.90) with the village in Malie being the one with higher entomological indicators and higher malaria incidence, and Upper Londolovit in the MIZ on the opposite position. On the other hand, there was no association between the entomological indicators and the prevalence of *Plasmodium spp* by qPCR. The results of this analysis can be seen in S3 Fig.

**Table 3.** Summary of vector and human metrics in each of the entomological sites.

| Geographic area | Village | Vector metrics |  |  | Human metrics |  |
| --- | --- | --- | --- | --- | --- | --- |
|  |  | SR | HBR | EIR | Prevalence | Incidence |
| Aniolam MIZ | Londolovit | 0.207 | 534.83 | 110.88 | 8.4% | 1.1 |
|  | Upper Londo | 0 | 31.96 | 0 | 8.5% | 1.1 |
|  | Kunaye 2 | 0 | 70.24 | 0 | 18.1% | 14.1 |
| Aniolam non-MIZ | Lienbel | 0.053 | 318.42 | 16.76 | 38.5% | 42.9 |
|  | Matakues | 0.077 | 716.8 | 55.14 | 32.8% | 20.3 |
|  | Sianius | 0.115 | 1664.43 | 190.92 | 20.0% | 18.9 |
| Malie Island | Malie (Sinambiet) | 0.063 | 8487.5 | 532.76 | 14.4% | 87.5 |
| Masahet Island | Masahet (Ton) | 0 | 4.57 | 0 | 9.5% | 9.7 |

Legend: Vector metrics such as human biting rate (in bites per person-year), sporozoite rate and entomological inoculation rate (percentage of infective bites per person-year), and human prevalence (percentage of infected individuals in the cross-sectional survey) and incidence of the month of December 2019 (number of cases per 1,000 inhabitants) for each of the villages with entomological surveillance. Abbreviations: EIR = entomological inoculation rate, HBR = Human Biting Rate, MIZ = mine-impacted zone, SR = sporozoite rate.

## DISCUSSION

This study characterized the human and vector determinants behind the different intensities of malaria transmission on the Lihir Islands of PNG, which are influenced by a gold mining operation. The MIZ of Aniolam, previously exhibiting lower prevalence (18), maintained a reduced burden with similar prevalence rates by microscopy. However, there has been a three-fold decrease in infection incidence in this area over the past decade. In contrast, the non-MIZ of Aniolam and Malie Island exhibited significantly higher infection rates, comparable to the highest endemicity areas in PNG such as the East Sepik (23). Notably, the north-western zone of Aniolam showed prevalence rates similar to those observed in heavily malaria-burdened countries in Africa (3). In this context, the study population, especially children, presented characteristics associated with high malaria exposure, such as moderate and severe anaemia, and splenomegaly (24). These clinical characteristics were more common outside the MIZ and were associated with carrying malaria parasites. Indeed, the strongest independent risk factor for carrying malaria parasites was the geographic location of the participants’ households.

As this study was conducted at the household level, we were able to demonstrate that, after geographic location, sharing a household with a positive individual was the strongest risk factor for malaria infection. This finding aligns with other studies conducted in Equatorial Guinea and Kenya (25, 26). Additionally, living in a traditional house was associated with a higher risk of infection, attributed to the open housing structures inherent in traditional dwellings, known to exacerbate malaria transmission (27). Of note, use of LLIN was low across the Lihir Islands, with only 37.2% of the participants sleeping under an LLIN the previous night. Although this percentage is higher than a previous study reporting a mere 13.6% LLIN use (19), LLINs remained insufficient as we did not observe any association between their use and the prevalence of infection.

*P. falciparum* and *P. vivax* were the most prevalent species identified in both the prevalence survey and the passive case detection analyses. Mixed infections were detected in a quarter of the positive cases when using qPCR, mainly due to the revealed *P. vivax* submicroscopic infections, and they were more frequent in children. These findings are consistent with reports from other regions of PNG and the Solomon Islands (28, 29). A previous study in PNG demonstrated that one of the factors contributing to this higher rate of *P. vivax* sub-microscopic infections in children is the occurrence of frequent relapses (30). A limitation of our study stems from assessing prevalence only once, hence not considering climatic patterns throughout the year. However, incidence of malaria in Lihir during 2019 showed no significant variations across months. Of note, we observed few discrepancies between microscopy and RDT in the prevalence survey. We could explain the additional *P. falciparum* infections detected by RDT due to persistence of HRP2 antigen during antimalarial treatment, while the non-*falciparum* infections missed by RDT were attributed to the lower sensitivity of this tool in detecting these species compared to microscopy (31). On the other hand, the incidence data showed a higher number of mixed infections when using RDT, probably due to some pLDH lines caused by high *P. falciparum* parasitaemia (32).

The entomology surveys confirmed a worryingly high outdoor and early biting feeding behaviour for the anopheline populations, particularly *An. faurauti*, consistent with previous findings in PNG (15). While a nationwide study showed no increase in the proportion of infective bites occurring before 10 pm (17), a subsequent study conducted in Madang Province highlighted the epidemiological significance of this earlier feeding behaviour by *An. farauti* (33). This discovery challenges the effectiveness of universal coverage with LLINs as the main, and often only, vector control strategy in our setting. People in PNG often spend a significant portion of their evenings and early nights engaged in outdoor activities without protection (27). The early biting behaviour of *An. farauti* has also been observed in the Solomon Islands (34), emphasizing the need for innovative vector control strategies targeting these species. Hence, our study supports that LLINs alone are insufficient to reduce malaria transmission in the Pacific; also, because many of the *P. vivax* infections are caused by relapses as explained above EIR are higher in the non-MIZ of Aniolam and Malie Island, areas of Lihir relying solely on LLINs, compared to the MIZ of Aniolam benefiting from alternative vector control interventions implemented by the mining operator.

Moreover, vector composition, proportion of larvae colonized sites, and transmission intensities were also different across the areas, aligning with malaria burden variations as seen in the correlation analysis, especially for the incidence data. Despite the open-pit mine and the abundant human dwellings in the MIZ of Aniolam, larvae colonized sites and HBR/EIR are low, possibly due to the specific larviciding conducted in this area (35), as well as better housing conditions (36). Conversely, Malie Island exhibits the highest HBR and EIR among the Lihir Islands, with most of the breeding sites found colonized by anophelines. The numerous mangroves located in this area may contribute to the highest density of *An. farauti*, as it is known to breed on brackish water and coastal streams (37). In contrast, the non-MIZ of Aniolam exhibits a diverse mosquito ecology with *An. punctulatus* as the main vector, which is considered more efficient (38) and potentially more anthropophilic (39). The environmental changes on Aniolam, including road extension, may explain the predominant presence of *An. punctulatus* outside the MIZ (40). Surprisingly, Masahet Island, relying solely on LLINs for vector control, had the lowest mosquito and larvae densities, along with a lower human burden. Masahet Island has reduced mangroves and swamps areas, and villages are coastal, with residents keeping livestock fenced several metres away from houses, unlike other areas of the Lihir Islands. A link between proximity of cattle to human dwellings and a higher risk of infection has been showed before (41). Hence, this distinct behaviour could explain the lower vector densities on Masahet Island, especially as the only vector identified there was *An. farauti*, a highly generalist mosquito (39).

Identifying human and vector factors for malaria risk serves for locating areas of concentrated malaria transmission and directing targeted measures to reduce the disease burden (42). Data on prevalence and incidence of *P. vivax* infections, particularly among children, underscore the need to enhance radical cure implementation to prevent relapses and maximize transmission reduction, as recommended by a cohort study in East Sepik (43). Focusing on the paediatric population is crucial in moderately to highly transmissive settings like Lihir, where interventions targeting transmission reduction would have the most significant impact (44). It may also be beneficial to explore specific measures for reducing intrahousehold transmission, such as reactive focal mass drug administration at the household and neighbouring levels; even though its effectiveness has been better demonstrated in low-endemicity settings (45).

Despite reported changes in *Anopheles* biting behaviours, it remains advisable to improve the usage of LLINs; however, the decreased bioefficacy of distributed nets in recent campaigns in PNG (46) encourages the need for regular monitoring of this strategy (47) and increase of funding, commitment, and innovative strategies (48). The evidence of lower malaria burden and reduced entomological metrics in Masahet and the MIZ of Aniolam supports the implementation of specific vector control strategies across the Lihir Islands. This could include measures like segregating livestock from the human population (49), exploring endectocidal treatments such as ivermectin for livestock (50), and expanding larviciding and environmental management to target breeding sites beyond the MIZ (51).

## CONCLUSION

The current study unveils unique transmission patterns within the Lihir Islands, emphasizing the necessity for customized strategies tailored to the specific characteristics of each area. However, considering the high permeability of these areas, both inside and outside the MIZ, it is crucial for the mine company to strengthen existing malaria control strategies and introduce innovative approaches across the whole group of Islands. This comprehensive analysis holds the potential to guide ongoing malaria control efforts and provide a roadmap for addressing similar challenges in New Ireland Province and other high-transmission coastal zones across the Western Pacific region.

## Data Availability

A dataset have been uploaded into the Supporting information files. This dataset have been modified to delete all information that could potentially identify or locate the study participants.

## ACKNOWLEDGEMENTS

We acknowledge all the participants in the study and the Lihirian communities for the acceptance and collaboration in this research. We thank Newcrest Mining Limited and Medicines for Malaria venture (MMV) for the research grant provided to PNG-IMR and ISGlobal as part of its collaborative agreement to support the Lihir Malaria Elimination Programme. We acknowledge support from the grant CEX2018-000806-S funded by MCIN/AEI/ 10.13039/501100011033, and support from the Generalitat de Catalunya through the CERCA Programme. CISM is supported by the Government of Mozambique and the Spanish Agency for International Development (AECID). BB is a Beatriu de Pinós postdoctoral fellow granted by the Government of Catalonia’s Secretariat for Universities and Research, and by Marie Sklodowska-Curie Actions COFUND Programme (BP3, 801370). SK was supported by a National Health and Medical Research Council Career Development Fellowship (GNT1141441).

## SUPPORTING MATERIAL CAPTIONS

**S1 Fig. Map of the Lihir Islands of Papua New Guinea**.

Located in the New Ireland Province, the Lihir Islands group is formed by three raised coral platform islands and two low coralline islets. The map shows the limits of the mine-impacted zone and the location of all the health facilities inside and outside this area.

**S2 Fig. Malaria cases diagnosed at the Lihirian health facilities**.

(A) Total number of malaria cases by age groups (unspecified group are adults’ patients without identified age); (B): *Plasmodium* species diagnosed by light microscopy and Rapid Diagnostic Test (RDT) in patients presented at the health facilities. For the RDT results, the health facilities recorded *P. falciparum* if the test showed a single line for histidine-rich protein 2 (HRP2), non *P. falciparum* (*P. vivax* in the figure) if the test showed a single line for *Plasmodium* lactate dehydrogenase (pLDH), and mixed infection if the test showed the two lines.

**S3 Fig. Correlation analysis between vector and human indicators in the Lihirian villages**.

(A) Correlation analysis between EIR and prevalence; (B) Correlation analysis between EIR and incidence; (C) Correlation analysis between HBR and prevalence; (D) Correlation analysis between HBR and incidence. HBR are expressed in bites per person-year, EIR are expressed in percentages, prevalence is expressed in percentage, and incidence is expressed in number of cases per 1,000 inhabitants. Abbreviations: CI = confidence interval, EIR = entomological inoculation rate, HBR = Human Biting Rate, MIZ = mine-impacted zone, p = correlation p-value, S = Spearman test value, rho = correlation coefficient.

**S1 Table. Characteristics of the participants included in the prevalence survey.**

Abbreviations: MIZ = mine-impacted zone, SD = Standard deviation. ^a^n = 2911 (0.1% missing), ^b^n = 1116 (0% missing, only for females with ≥ 16 years-old), ^c^n = 805 (0% missing, only for participants between 5-18 years-old), ^d^n = 2,912 (0.1 % missing), ^e^n = 2,884 (1.0 % missing), ^f^n = 1486 (49.0% missing), ^e^n = 1877 (35.6% missing).

**S2 Table. Malaria incidence risks in the Lihir Islands by age groups and geographic areas.**

Abbreviations: CI (Confidence Interval), IR (Incidence Risk), IRR (Incidence Risk Ratio), MIZ (Mine-impacted zone). Binomial regression model was used to estimate IRR with 95% CI and p-values.

**S3 Table. Type of habitats where anophelines were found in the Lihir Islands**.

Abbreviations: CI (Confidence Interval), N (number), Prop (proportion).

**S1 File. Supplementary methods**.

